# Optimal size of sample pooling for RNA pool testing: an avant-garde for scaling up SARS CoV 2 testing

**DOI:** 10.1101/2020.06.11.20128793

**Authors:** Khodare Arvind, Padhi Abhishek, Gupta Ekta, Agarwal Reshu, Dubey Shantanu, SK Sarin

## Abstract

**Introduction:** Timely diagnosis is essential for the containment of the disease and breaks in the chain of transmission of SARS-CoV-2. The present situation demands countries to scale up their testing and design innovative strategies to conserve diagnostic kits and reagents. The pooling of samples saves time, manpower, and most importantly diagnostic kits and reagents. In the present study, we tried to define the pool size that could be applied with acceptable confidence for testing.

**Material and methods:** We used repeatedly tested positive clinical sample elutes having different levels of SARS CoV 2 RNA and negative sample elutes to prepare seven series of 11 pools each, having pool sizes ranging from 2 to 48 samples to estimate the optimal pool size. Each pool had one positive sample elute in different compositions. All the pools were tested by SARS CoV 2 RT-qPCR.

**Results:** Out of the 77 pools, only 53 (68.8%) were found positive. The sensitivity of pools of 2 to 48 samples was decreased from 100% (95% CL; 98.4-100) to 41.41% (95% CL; 34.9-48.1). The maximum size of the pool with acceptable sensitivity (>95%) was found to be of 6 samples. For the pool size of 6 samples, the sensitivity was 97.8% and the efficiency of pooling was 0.38.

**Conclusion:** The pooling of samples is a practical way for scaling up testing and ultimately containing the further spread of the COVID-19 pandemic.

## Introduction

What started as a cluster of pneumonia cases in Wuhan city of China has become now a full-blown pandemic, the first of its kind due to the coronavirus ^[1]^. This pandemic popularly known as COVID-19 (coronavirus disease 2019) is caused by SARS-CoV-2 (severe acute respiratory syndrome coronavirus-2)^[2]^. As of 3^rd^ June 2020, COVID-19 had globally caused 6, 287, 771 confirmed cases and 379, 941 deaths affecting over 200 countries ^[3]^. Timely diagnosis is essential for the containment of the disease and breaks in the chain of transmission of SARS-CoV-2.

The laboratory diagnosis of SARS-CoV-2 is based primarily on nucleic acid amplification test (NAAT) like real-time reverse transcriptase PCR (RT-qPCR). As the number of COVID-19 cases is increasing every day the availability of diagnostic kits and reagents has emerged as a major bottleneck in the laboratory testing of SARS-CoV-2 ^[4]^. The currently available testing strategies mainly focus on symptomatic individuals. But detecting the carrier or asymptomatic individuals holds the key in containing the spread of the infection into the community. Earlier the infected person is identified, sooner the spread of the infection can be contained and the surveillance machinery can be activated for contact tracing and ultimately break in the chain of transmission of the virus^[5]^. Densely and heavily populated countries like India and other developing countries have abysmally low test rates per capita in the world^[6]^ and the number of cases is expected to shoot up post lockdown. So the present situation demands countries to scale up their testing and design innovative strategies to conserve diagnostic kits and reagents.

The pooling of diagnostic tests, which has previously been applied in other infectious diseases^[7][8]^, is one such method where samples are mixed and tested at a single pool, and subsequent individual tests are made only if the pool tests positive. Pooling saves time, manpower, and most importantly diagnostic kits and reagents.

In the present study we tried to define the pool size that could be applicable to a particular population with acceptable confidence and avoiding drop of positive cases which is a major risk of pool testing.

## Material and methods

### Sample collection

Combined nasopharyngeal and oropharyngeal swabs were collected and transported in viral transport media(VTM) maintaining the proper cold chain and sent to the virology laboratory of the Institute of Liver and biliary sciences, New Delhi, India. A volume of 200 microlitres (µl) of the sample was further processed for viral nucleic acid extraction by Qiasymphony DSP Virus/ Pathogen mini kit (Qiagen GmbH, Germany) as per the manufacturer’s protocol in elutes of 60 µl each^[9]^. Each sample was subjected to the addition of 10 µl of spiked extraction control (EAC) at the time of extraction itself, to check the validity of the extraction procedure.

### Performance of RT-qPCR in the laboratory

The 5 µl of the extracted RNA elute/sample was subjected to RT-qPCR for the qualitative detection of SARS-CoV-2 RNA utilizing with AgPath-IDTM One-Step RT-PCR Reagents (Thermo Fisher Scientific) using an Applied biosystem (ABI) 7500 Real-Time PCR system (Thermo Fisher Scientific) and LightMix® SarbecoV E-gene (TIB MOLBIOL). Reactions were heated to 55°C for 5 minutes for reverse transcription, denatured in 95°C for 5 minutes, and then 45 cycles of amplification were carried out for 95°C for 5 seconds and 60°C for 15 seconds using FAM parameter for E gene. This assay targets the detection of the E gene for SARS as well as nCoV-2. The primer details are given below.

Primers and probes for the RT-qPCR^[10]^

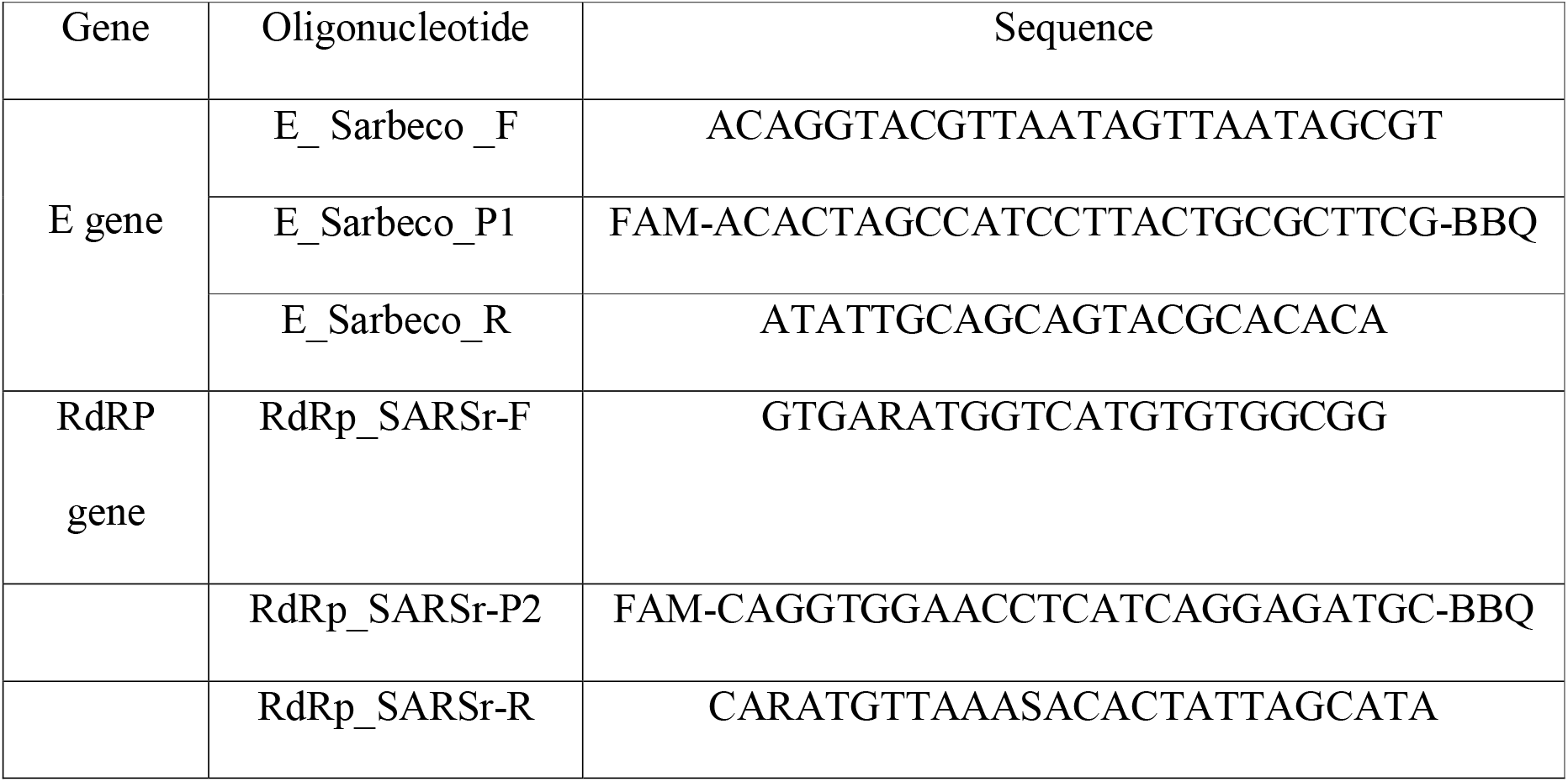

FAM: 6-carboxyfluorescein; BBQ: blackberry quencher.

All samples that were screened positive for the E gene were confirmed by the performance of RT-qPCR for the detection of specific RdRp gene of SARS-CoV-2 using LightMix® Modular SARS-CoV-2 RdRP (TIB MOLBIOL) using similar PCR conditions as described above.

### Scheme of pooling of RNA elutes

Arbitrarily 7 twice tested SARS CoV 2 E & RdRp gene positive RNA elutes with varying cycle threshold values and 48 twice tested negative elutes for SARS CoV 2 E & RdRp gene tested by utilizing AgPath-IDTM One-Step RT-PCR Reagents were selected. A total of 48 negative sample elutes were used to make 8 series of 11 pools each (total of 77 pools) in an equal volume (3µl each) of 2, 4, 6, 8, 10, 12, 16, 20, 24, 32, and 48 sample elutes. Each of the 11 pools was mixed with 1 positive elute to make its 7 dilutions series of 1:2, 1:4, 1:6, 1:8, 1:10, 1:12, 1:16, 1:20, 1:24, 1:32, and 1:48 dilutions. A 5µl of positive sample elute was mixed with the 5µl of a pool of 2 samples to make 1:2 dilution then this 1:2 mixture was serially double diluted to make 1:4, 1:8, 1:16, and 1:32 dilations using the pools of 4, 8, 18, and 32 sample elutes, respectively. A 3µl of positive sample elute was mixed with the 15µl of a pool of 6 samples to prepare 1:6 dilution of positive sample then this mixture was used to prepare serial double dilution of 1:12, 1:24, and 1:48 dilution using pools of 12, 24, and 48 samples, respectively. To prepare a dilution of 1:10 the 3µl volume of positive sample was mixed with 27µl of a pool of 10 samples then it was double diluted with a pool of 20 samples to make 1:20 dilution of a positive sample. The cycle threshold of all the 7 positive samples used for serial dilution was 25, 29, 31, 33, 35, 38, and 39. A separate 8^th^ dilution series of 11 pools were tested by adding PCR grade water instead of positive sample elutes which acted as a control. 5µl of elute from the pool was used for RT-qPCR. Different 48 negative elutes were used to prepare the pools, to determine the effect of the individual negative sample (to establish the heterogeneity) in detection of a positive sample in the pool.

The frequency of Ct values distribution of individually tested positive samples reported from our laboratory during this pandemic was estimated and were categorized into ≤ 25, >25 to 29, >29 to 31, >31 to 33, >33 to 35, >35 to 38 and >38 to 40 ranges. The frequency of positive pools falling in these ranges was used to calculate the sensitivity of the pooling of sample elutes for RT-qPCR.

Known positive and negative sample elutes were used for making of pools, but for calculation, it was considered that if following a pooling algorithm after obtaining results of pool testing, the same previously obtained results will get after individual testing of samples. The result of individually tested samples was considered the gold standard for the calculation of sensitivity, specificity, negative predictive value (NPV), and positive predictive value.

## Results

In this study, clinical samples previously established as positive for the SARS CoV-2 virus were chosen to determine whether they can detectable when their elutes mixed with negative samples elutes in different dilutions. Positive sample elutes for SARS CoV-2 with a different range of Ct values of E gene target were chosen, serially diluted with negative sample elutes, and RT-qPCR was performed.

Overall 88 pools were made which included 77 pools having one positive sample elute in different concentrations and 11 pools having no positive sample elutes. Out of the 77 pools of up to 48 times serial dilutions of positive samples, only 53 (68.8%) were found positive. (Table-1)

**Table 1.**
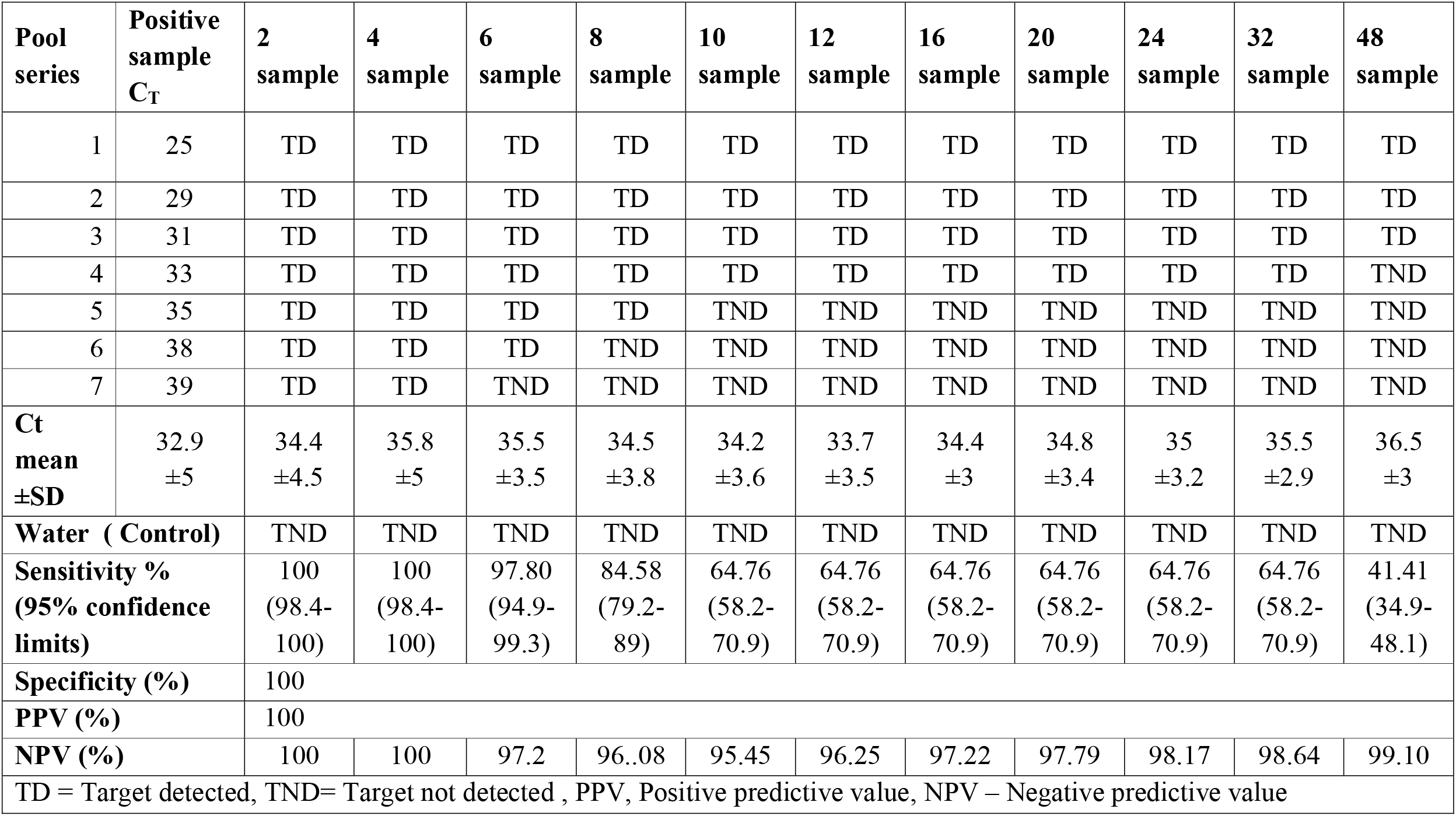
Results, mean Ct values and clinical performance of pool testing.

| **Pool series** | **Positive sample C<sub>T</sub>** | **2 sample** | **4 sample** | **6 sample** | **8 sample** | **10 sample** | **12 sample** | **16 sample** | **20 sample** | **24 sample** | **32 sample** | **48 sample** |
| --- | --- | --- | --- | --- | --- | --- | --- | --- | --- | --- | --- | --- |
| 1 | 25 | TD | TD | TD | TD | TD | TD | TD | TD | TD | TD | TD |
| 2 | 29 | TD | TD | TD | TD | TD | TD | TD | TD | TD | TD | TD |
| 3 | 31 | TD | TD | TD | TD | TD | TD | TD | TD | TD | TD | TD |
| 4 | 33 | TD | TD | TD | TD | TD | TD | TD | TD | TD | TD | TND |
| 5 | 35 | TD | TD | TD | TD | TND | TND | TND | TND | TND | TND | TND |
| 6 | 38 | TD | TD | TD | TND | TND | TND | TND | TND | TND | TND | TND |
| 7 | 39 | TD | TD | TND | TND | TND | TND | TND | TND | TND | TND | TND |
| **Ct mean ±SD** | 32.9 ±5 | 34.4 ±4.5 | 35.8 ±5 | 35.5 ±3.5 | 34.5 ±3.8 | 34.2 ±3.6 | 33.7 ±3.5 | 34.4 ±3 | 34.8 ±3.4 | 35 ±3.2 | 35.5 ±2.9 | 36.5 ±3 |
| **Water ( Control)** |  | TND | TND | TND | TND | TND | TND | TND | TND | TND | TND | TND |
| **Sensitivity % (95% confidence limits)** |  | 100 (98.4-100) | 100 (98.4-100) | 97.80 (94.9-99.3) | 84.58 (79.2-89) | 64.76 (58.2-70.9) | 64.76 (58.2-70.9) | 64.76 (58.2-70.9) | 64.76 (58.2-70.9) | 64.76 (58.2-70.9) | 64.76 (58.2-70.9) | 41.41 (34.9-48.1) |
| **Specificity (%)** |  | 100 |  |  |  |  |  |  |  |  |  |  |
| **PPV (%)** |  | 100 |  |  |  |  |  |  |  |  |  |  |
| **NPV (%)** |  | 100 | 100 | 97.2 | 96..08 | 95.45 | 96.25 | 97.22 | 97.79 | 98.17 | 98.64 | 99.10 |
| TD = Target detected, TND= Target not detected , PPV, Positive predictive value, NPV – Negative predictive value |  |  |  |  |  |  |  |  |  |  |  |  |

Positive samples with Ct values ranging from 25 to 31 were detected in pools till 1: 48 dilution, while pools containing samples of Ct value 33 were detected till 1:32 dilution. Pools with higher Ct values became negative in lower dilutions. Sample with Ct values of 35, 38, and 39 was detected till 1:8, 1:6 and 1:4 dilutions respectively. As the Ct value of an individual positive sample in the pool was increasing, the probability of detection of the positive pool was decreasing and as the size of the pool increased, Ct was attained later, as expected due to the dilution effect. (Table-1, Figure-2)

The frequency of Ct values (E gene only) distribution of individually tested samples was derived from 227 SARS CoV 2 E & RdRp gene positive samples detected from 1^st^ of March to 30^th^ April 2020 using the same PCR reagents used for pool testing. (Table-2)

### Calculation of sensitivity of pool testing

Sensitivity of pool testing was defined as the probability that a true positive individual sample will be declared positive. The sensitivity of pooling was calculated by taking into consideration of the distribution of a particular Ct value in results obtained during routine diagnostic testing of individual samples received. We defined 7 ranges of Ct values and for pooling, we took samples with Ct value equal to the upper limit of the range as a more conservative approach. (Table 2, Figure-1) All the pools of 6 samples having positive samples of Ct values 25-38 was tested positive and the Ct values of 97.8% of individually positive samples during this pandemic were distributed within Ct value range of 25-38, so the calculated sensitivity of the pool of 6 samples was 97.8% (95% CL: 94.9-99.3). Similarly, the sensitivity of other pool sizes was calculated and shown in table 1. The sensitivity of pool testing decreased as the pool size increased and also as the Ct value of the positive sample in the pool increased. The acceptable sensitivity (> 95%) of pool testing was found for pooling of up to 6 samples. (Table-2, Figure-3)

**Figure 1.**
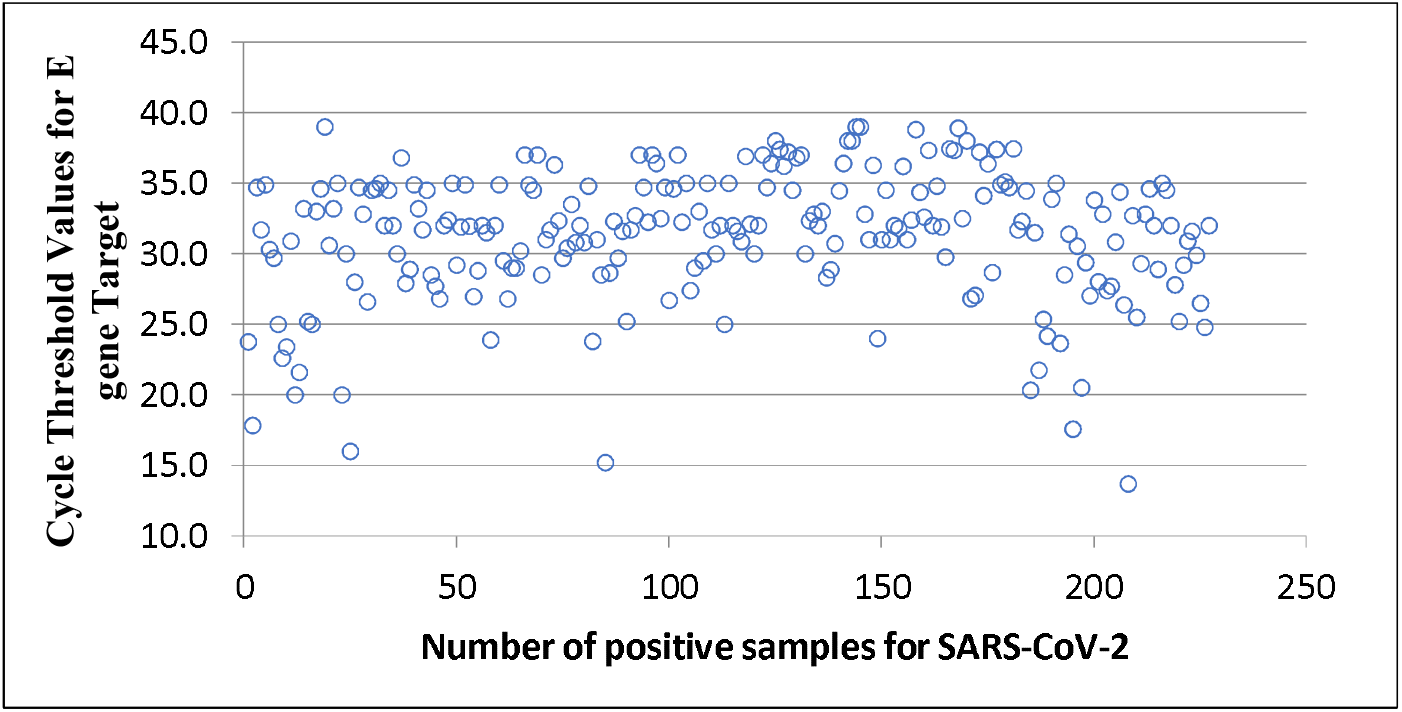
Cycle threshold values of 277 samples tested positive for E gene target of SARS-CoV 2.

**Table 2.** Distribution of Ct value in positive samples received from 1^st^ March to 30^th^ April 2020 and Ct values of positive samples used in pooling & sensitivity of pools.

| S. No. | Distribution of Ct value in tested positive samples, n=227 |  |  | Ct value of the Positive sample in pool | Maximum pool size detected positive | Sensitivity (95% CL) of pool testing |
| --- | --- | --- | --- | --- | --- | --- |
|  | Ct range | Number | % |  |  |  |
| 1. | < 25 | 23 | 10.1 | 25 | 48 | 41.41<br>(34.9-48.1) |
| 2. | 25-29 | 37 | 16.3 | 29 | 48 |  |
| 3. | 29-31 | 34 | 15.0 | 31 | 48 |  |
| 4. | 31-33 | 53 | 23.3 | 33 | 32 | 64.76<br>(58.2-70.9) |
| 5. | 33-35 | 45 | 19.8 | 35 | 8 | 84.58<br>(79.2-89) |
| 6. | 35-38 | 30 | 13.2 | 38 | 6 | 97.80<br>(94.9-99.3) |
| 7. | > 38 | 5 | 2.2 | 39 | 4 | 100<br>(98.4-100) |
| Median (IQR) | 32<br>(34.7-29.0) |  |  | 33<br>(36.5 – 30) |  |  |
| CL- Confidence limit, IQR- Inter quartile range |  |  |  |  |  |  |

### Calculation of specificity of pool testing

Specificity of pool testing was defined as the ability of the pooling algorithm to correctly identify those without the infection. For the calculation of specificity of pooling a total of 11 pools of 2, 4, 6, 8, 10, 12, 16, 20, 24, 32 and 48 SARS CoV 2 E & RdRp gene negative samples elutes were tested. All the 11 pools tested negative and false positive results were nil, so the specificity of pooling of sample elutes for SARS-CoV-2 RT-qPCR was 100%.

### NPV of pool testing

The NPV was defined as the probability that an individual specimen identified as negative at the end of a pooling algorithm was truly negative. The overall negative predictive value was found to be 97.8%. The NPV for a pool size of 2 and 4 was 100% and found no false negative results but for the pool sizes of 6, 8, and 10, NPV gradually decreased from 97.2 % to 95.45 % due to gradual increase in false negative results. For pool sizes from 12 to 48, NPV was progressively increased due to stable false negative results but increasing pool size.

### PPV of pool testing

The PPV was defined as the probability that a specimen identified as positive at the end of a pooling algorithm was truly positive. The PPV of pooling was 100% and no false positive results were found.

### Efficiency of sample pooling

The efficiency of a pooling algorithm was defined as the expected number of NAATs required per individual specimen evaluated. An efficiency of sample pooling was calculated 0.38 using the online calculator considering the two-stage Dorfman mini-pool strategy of pool testing with the conservative predictions of a sensitivity of 95%, a specificity of 99%, 4% prevalence of SARS-CoV-2 infection and pool size of 6 samples. ((http://www.bios.unc.edu/∼mhudgens/optimal.pooling.b.htm)

### Calculation of optimum pool size

Most positive pools reach the threshold at a later Ct as they are further diluted. A pool size of 6 samples having a sensitivity of 97.8% and NPV of 97.2% was considered acceptable for the pooling of sample elutes for RT-qPCR.

## Discussion

While individual level nucleic acid amplification testing (NAAT) remains the gold standard for the diagnosis of COVID-19 infection, a limited supply of diagnostic kits and reagents remains the major bottleneck in expediting testing of COVID-19 in the community especially during surge testing^[11]^. Furthermore, mass testing of samples should be done to control the COVID-19 pandemic at the earliest ^[12]^. Hence a testing method where a large number of samples can be tested and consuming minimal testing kits and reagent is the need of the hour. In this study effect of pool size on detection of SARS CoV 2 and its accuracy in the population was assessed. We found that a single clinical sample with SARS-CoV-2 RNA can be consistently detected in a pool of a maximum of four samples. The Ct values of individually tested RNA elute and that of the pooled group were in sync. As evident from figure 2 pooling in this pattern leads to a gradual increase in the threshold cycle of the pooled group. Furthermore, additional amplification cycles could lower the detection limit allowing better detection of pools having samples of higher Ct values ^[13]^.

**Figure 2.**
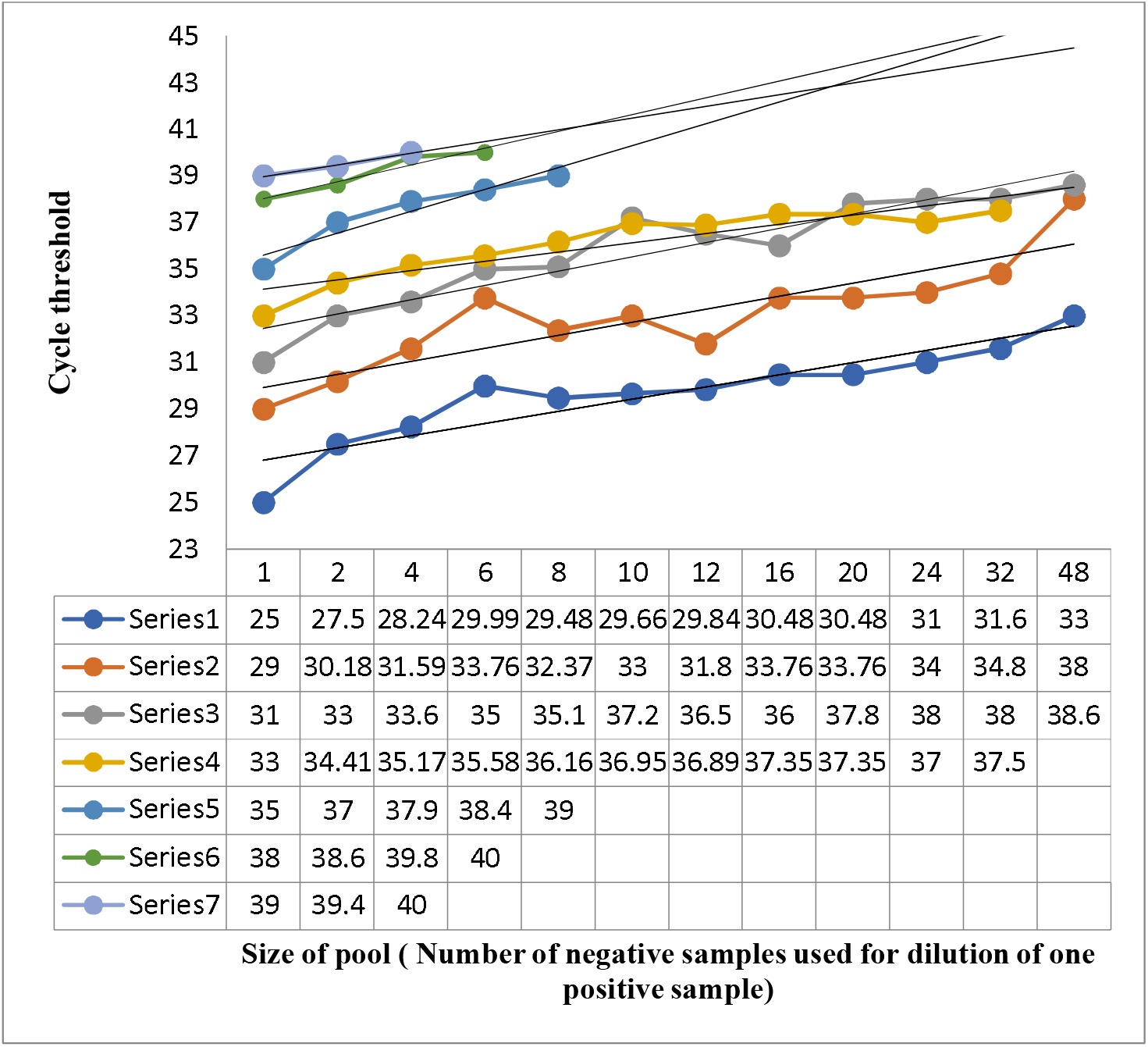
Ct values of pools having a positive sample of different Ct and trend of increase of Ct value as the pool size increases.

**Figure 3.**
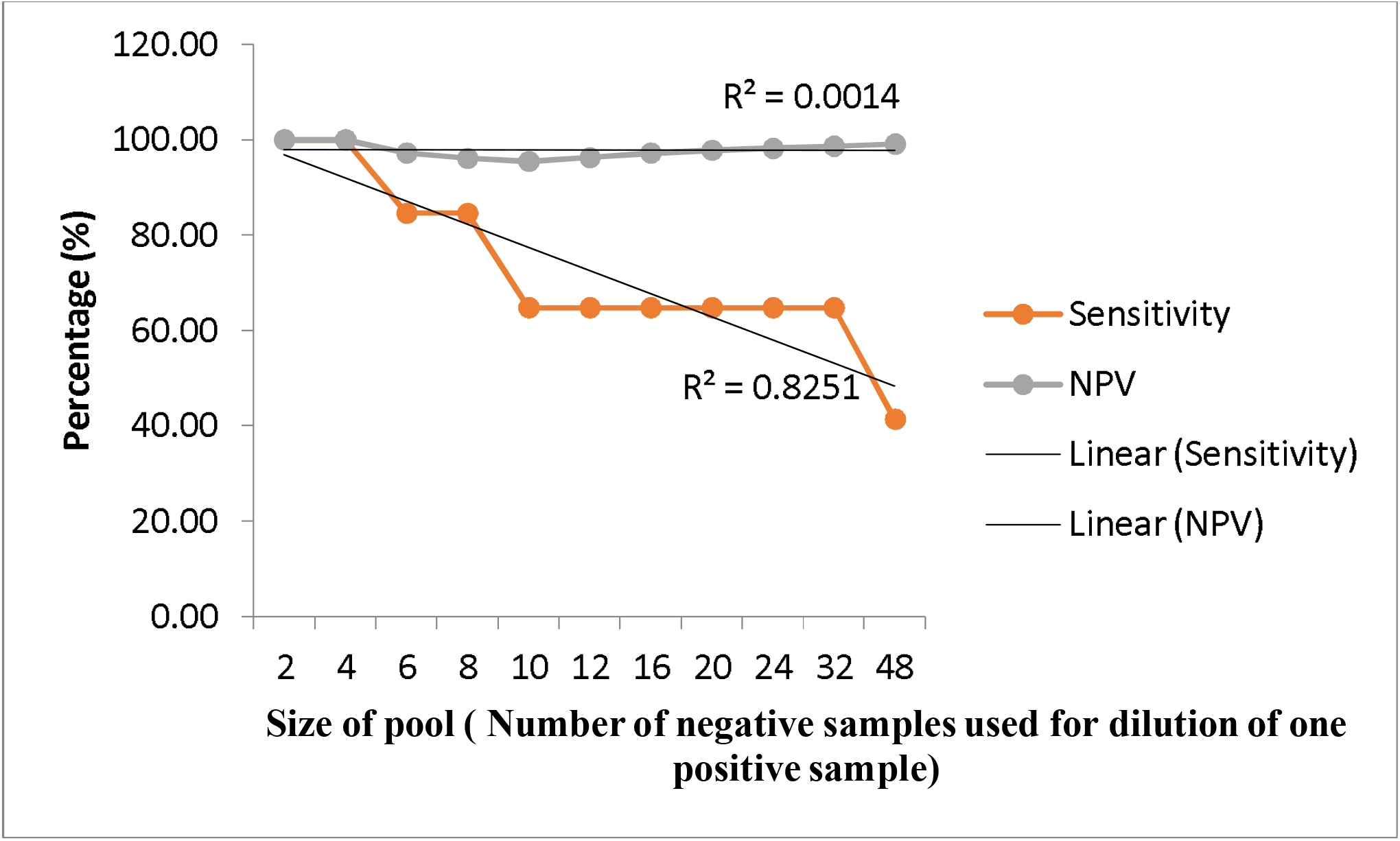
Sensitivity and NPV of pools of different sizes. Figure 3 shows the trend of sensitivity and NPV of pooling as the pool size increased.

The pooling of samples is essentially important in monitoring the infection in cohesive groups such as quarantine facilities, health care workers, community surveillance, and diagnosing the asymptomatic cases. The infection load in these groups may be low but even a single positive case amongst such groups can activate the surveillance system and quarantine the affected group and prevent the further spread in the community. The pooling is better suited for low prevalence populations and it could fasten the testing over a very short time that would be able to identify new hot spot areas for infection before becoming condition worst. The pooling of samples can also be done prior to RNA extraction that is at the time of sample collection, putting the nasopharyngeal/ oropharyngeal swabs into a common viral transport media (VTM) and after sample collection by pooling of VTM. By doing so, the major bottleneck of RNA extraction can be removed.

The pooling of samples leads to the dilution of nucleic acid present in the pool leading to a decrease in the sensitivity. Hence the pool size should be optimized for the agent to be detected and assay to be used so that even low positive samples would not be missed in the pool testing. Gupta E et al. showed that the increase in the cellular material, including nucleic acid, due to the pooling of multiple samples did not affect the detection of the SARS-CoV-2 virus by RT-qPCR.^[14]^ The size of the pooling of samples should be such that it should give a positive result if it has at least one positive sample that has the lowest amount of viral nucleic acid so that the sensitivity does not get compromised. We determined the pool size by considering of amount of viral nucleic acid (as Ct value is usually proportional to the amount of nucleic acid) that is more prevalent in the community during this pandemic. A study by Arons et al. has shown that SARS-CoV-2 N1 targeted median Ct values for the four symptom status groups were similar (asymptomatic residents, 25.5; pre symptomatic residents, 23.1; residents with atypical symptoms, 24.2; and residents with typical symptoms, 24.8) ^[15]^. The median Ct value for samples tested positive in this pandemic was 32(34.7-29.0). 97.8% of samples had Ct value of <38 for the 45 cycle protocol of RT-qPCR and all the pools having a positive sample of Ct value <38 were tested positive up to the pool size of 6 samples, so we determined the optimal pool size of 6 for pool testing of SARS-CoV-2 RT-qPCR. The sensitivity (97.8%) and NPP (97.22%) value of the pool of 6 samples was taken to consider it optimal pool size. The samples that had a Ct value of >38 were only 2.2%. The clinical significance of such low positive samples could not be determined because of the unavailability of gold standard viral culture methods.

The efficiency of individual testing is 1, and efficiency of less than 1 indicates that the pooling algorithm will require fewer tests on average than individual testing. By calculating on online calculator we found the efficacy of pool testing of 6 samples 0.38 i.e. for one sample testing only 0.38 test reagents are required (>95% sensitivity of the test, a prevalence rate of infection 4% and two-stage Dorfman mini-pool strategy used for calculation). Many pool testing strategies for pooling are known. The two-stage Dorfman mini-pool strategy is one of them which is more convenient for pools of small sizes. In the first stage of this strategy, the master pool comprising all specimens is tested; if the master pool tests positive, all component individual specimens are tested (the second of the two stages) ^[16]^.

## Conclusion

As the COVID-19 pandemic continues spreading there is tremendous stress on logistics. Hence pooling of samples is a practical way for scaling up testing and ultimately containing the further spread of the COVID-19 pandemic. This study suggests that a pool size of 6 samples would be optimal with acceptable confidence.

## Data Availability

All the data has been mentioned in the manuscript.

## Acknowledgements

We acknowledge all the technical staff involved in testing and ICMR for providing the Primers and probes for the conduct of RT-q PCR.

